# CandyCodes: Simple universally unique edible identifiers for confirming the authenticity of pharmaceuticals

**DOI:** 10.1101/2021.07.30.21261395

**Authors:** William H. Grover

## Abstract

Counterfeit or substandard medicines adversely affect the health of millions of people and cost an estimated $200 billion USD annually. Their burden is greatest in developing countries, where the World Health Organization estimates that one in ten medical products are fake. In this work, I describe a simple addition to the existing drug manufacturing process that imparts an edible universally unique physical identifier to each pill, tablet, capsule, caplet, etc. This technique uses nonpareils (also called sprinkles and “hundreds and thousands”), tiny inexpensive multicolor candy spheres that are normally added to other candies or desserts as decorations. If nonpareils are applied at random to a pill immediately after manufacture, the specific pattern they form is unlikely to ever be repeated by random chance; this means that the pattern (or “CandyCode”) can be used to uniquely identify the pill and distinguish it from all other pills. By taking a photograph of each CandyCoded pill after manufacture and recording the location and color of each nonpareil, a manufacturer can construct a database containing the CandyCodes of all known-authentic pills they produce. A consumer can then simply use a cellphone to photograph a pill and transfer its image to the manufacturer’s server, which determines whether the pill’s CandyCode matches a known-good CandyCode in their database (meaning that the pill is authentic) or does not have a match in the database (in which case the consumer is warned that the pill may be counterfeit and should not be consumed). To demonstrate the feasibility of using random particles as universal identifiers, I performed a series of experiments using both real CandyCodes (on commercially produced chocolate candies) and simulated CandyCodes (generated by software). I also developed a simple method for converting a CandyCode photo to a set of strings for convenient storage and retrieval in a database. Even after subjecting CandyCodes to rough handling to simulate shipping conditions, the CandyCodes were still easily verifiable using a cellphone camera. A manufacturer could produce at least 10^17^ CandyCoded pills—41 million for each person on Earth—and still be able to uniquely identify each CandyCode. By providing universally-unique IDs that are easy to manufacture but hard to counterfeit, require no alteration of the existing drug formation and minimal alteration of the manufacturing process, and need only a cameraphone for verification, CandyCodes could play an important role in the fight against fraud in pharmaceuticals and many other products.

## 1 Introduction

Every year, an estimated $200 billion USD is wasted on medicines that are counterfeit or substandard. These fake medicines often appear authentic, but they may not contain the specified amounts of active ingredients, or may contain inert (or dangerous) additional ingredients. The problem of adulterated medicines is most acute in the developing world, where the World Health Organization estimates that one in ten medical products are counterfeit or adulterated [1]. When criminals copy or reuse the packaging of authentic brands, fake medicines devalue and erode public trust in those brands. Consequently, legitimate drug makers are keenly interested in defending their brands and protecting the drug distribution chain connecting their factories to consumers.

Unique identifiers can play an important role in defending the pharmaceutical distribution chain and fighting counterfeit medications. For example, legitimate manufacturers already print unique lot numbers on medication bottles; concerned consumers could contact the manufacturer and confirm that a given bottle’s number corresponds to a known-good lot. However, criminals could thwart this defense by reusing authentic packaging or printing known-good lot numbers on their own fake packaging. Additionally, somewhere along the distribution chain, a criminal could obtain an authentic bottle of medicine, replace the contents with a cheaper substitute, and re-seal the bottle. Finally, since pharmacies commonly dispense medicines in generic containers, an unscrupulous pharmacy could dispense fraudulent medication in bottles with no traceable lot numbers. In each of these scenarios, a unique lot number on drug packaging cannot combat counterfeiting.

The ideal identifier for combatting drug fraud would be *part of the drug product itself*, permanently linked to the medicine at every step from manufacturing all the way to the consumer. A consumer could use this on-drug identifier to confirm the authenticity of a drug immediately before taking it. Various versions of on-drug identifiers have been proposed, but each version has short-comings that have limited its widespread adoption. For example, one technique uses indentations on a drug capsule’s surface to encode an ID, but decoding the ID requires the use of a specialized reader [2]. Other techniques use fluorescent ink [3], lithographic patterning [4], or molding [5] to include a QR code on a pill or capsule; these IDs can be read using a standard smartphone camera, but integrating these codes into drug products would require a significant modification of the drug manufacturing process. Still other approaches print IDs onto film-based [6] or paper-based [7] formulations of a drug, but this requires a significant reformulation of the drug product. In summary, on-drug identifiers could play a role in combatting fraudulent pharmaceuticals, but problems with existing approaches to on-drug IDs have precluded their widespread adoption.

Researchers have also shown that *random physical patterns* can serve as unique IDs. For example, the “physical one-way functions” developed by Ravikanth Pappu *et al*. [8] consist of small glass particles randomly distributed in a transparent matrix; when laser light shines through the matrix, the optical speckle fluctuations in the transmitted light are unique to that particle pattern and are unlikely to ever be duplicated by chance. Since these “physical one-way functions” are easy to make but very difficult to duplicate, Pappu *et al*. recognized that they could be used as authentication tokens in cryptographic applications. Later examples of these “physical unclonable functions” or PUFs have used random patterns of molecules, nanotubes, nanoparticles, and other objects to uniquely identify objects [9]. Most recently, Jung Woo Leem *et al*. [10] demonstrated that fluorescent silk microparticles from genetically engineered silkworms can be distributed randomly in a silk film, then a piece of this silk can be attached to a pill. By imaging the silk-tagged pill using a fluorescence microscope, the microparticle pattern can be read and used to uniquely ID the pill. While these demonstrations confirm that random patterns can be used to uniquely identify a pill, they also require specialized equipment for reading the pill’s ID, equipment like lasers and fluorescence microscopes which are obviously not available to most consumers. This makes existing methods unsuitable for at-home use by an individual wishing to verify the authenticity of a pill before consuming it.

In this work, I describe a simple addition to the standard pharmaceutical manufacturing process that imparts to each pill a universally unique identifier that can be read using an ordinary cameraphone. This method was inspired by the chocolate candies coated in tiny multicolored candy “nonpareils” shown in Figure 1. If enough multicolored nonpareils are applied at random to the chocolate, the pattern the nonpareils make can be considered universally unique and unlikely to ever be replicated by chance on another chocolate. Moreover, these patterns are simple to create (simply apply nonpareils at random), but it would be very difficult and time-consuming to duplicate an existing pattern by hand. These properties make multicolored nonpareil coatings (or “CandyCodes”) highly suitable for use as on-drug identifiers for combatting fraudulent pharmaceuticals. As a proof-of-concept, I created both real and simulated CandyCodes and tested their suitability as universally-unique identifiers. My findings suggest that CandyCodes could be a useful tool in the fight against fraudulent medicines.

**Figure 1:**
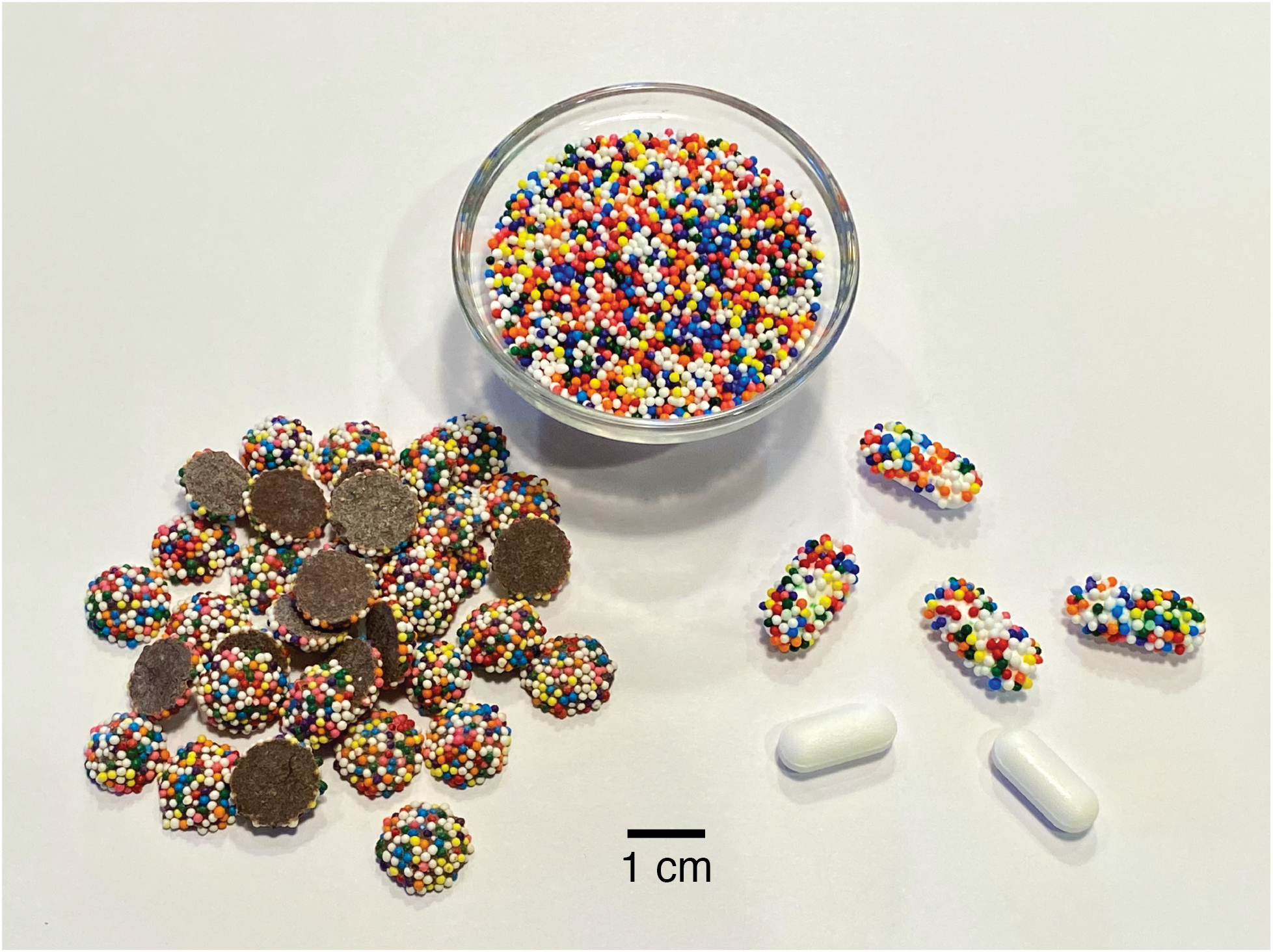
If enough multicolored particles (like these candy “nonpareils,” top) are randomly attached to objects, the odds of two objects having the same pattern of particles are essentially zero. This means that every candy-coated chocolate (left) is likely unique and one-of-a-kind in the entire world. By attaching the same multicolored candies to pharmaceutical products (like the acetaminophen/paracetamol caplets at right), each pill receives an edible universally unique identifier—a “CandyCode”—that a consumer can use to confirm the authenticity of the product.

## 2 Results

The overall process of using CandyCodes to guarantee the authenticity of a pharmaceutical product is summarized in Figure 2. Each pill is coated in random multicolored candy nonpareils and photographed before leaving the manufacturer. These photographs can be stored in a database as is, or they can be converted to a more-easily-searchable format (a method for converting a CandyCode photo to a set of text-based strings is described later in this work). Either way, a database is created containing CandyCode patterns belonging to known-good pills. When a consumer receives a pharmaceutical product and wishes to check the authenticity of a pill, they merely snap a photograph of the pill using a cellphone and upload it to the manufacturer’s server. The suspect pill’s CandyCode is then compared to the database of known-good CandyCodes. If a close match is found, the consumer is informed that the pill is authentic, but if no matching pattern is found, the consumer is warned that the drug product is not authentic and should be discarded. CandyCodes can be added to existing drug products without having to reformulate the product or significantly alter the manufacturing process, they need no specialized equipment to read, and they would be extremely difficult and time-consuming to duplicate in an attempt to circumvent the security they provide.

**Figure 2:**
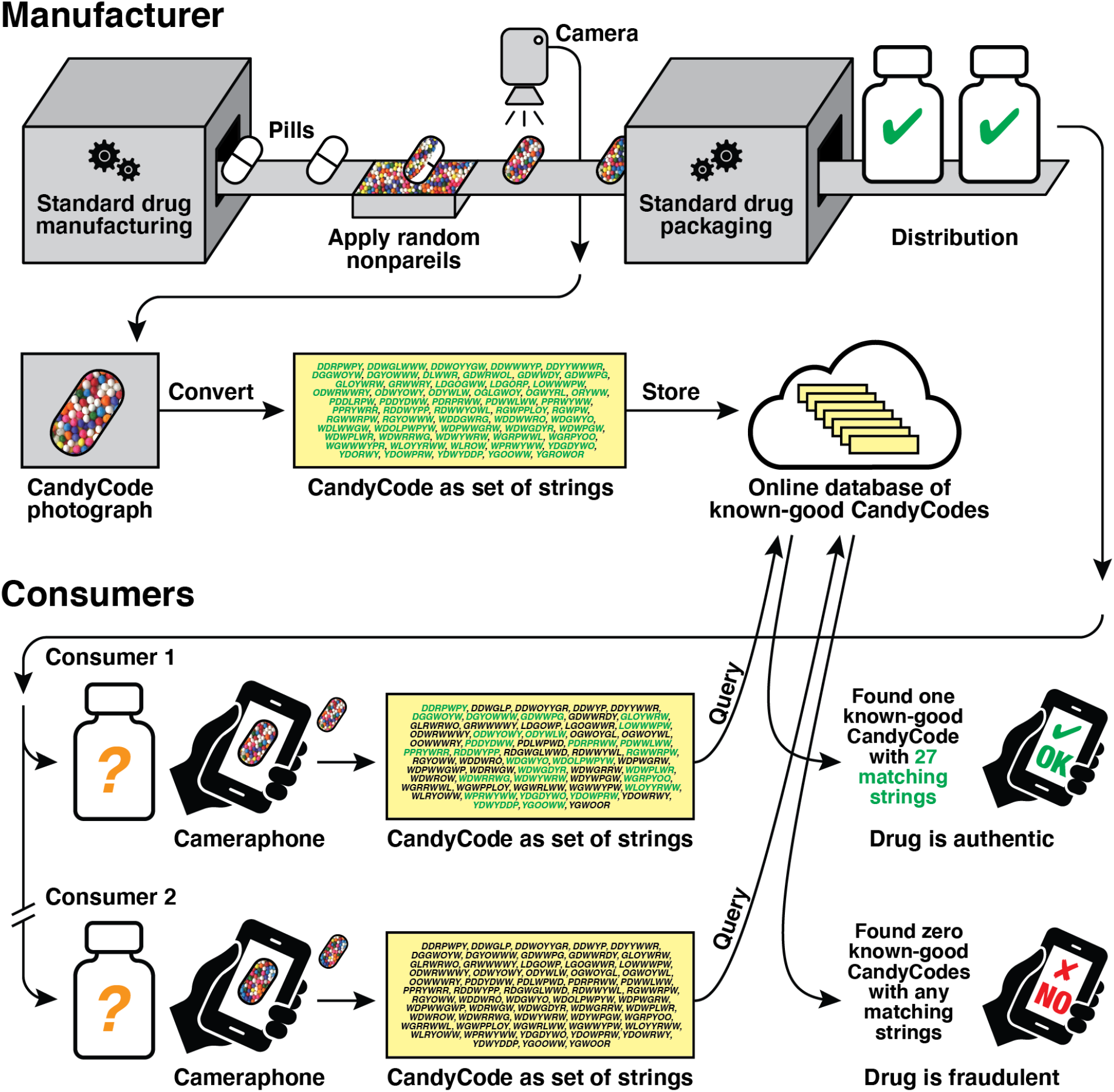
Using CandyCodes to authenticate pharmaceuticals. By randomly affixing a number of distinguishable edible particles (in this case, multicolored nonpareil candies) to each pill after manufacturing, a pharmaceutical company gives each pill a universally-unique “CandyCode.” Each CandyCoded pill is photographed before packaging and distribution, and the pattern of particles in each photo is recorded (in this example, the manufacturer converts each photo into a set of text strings using a process detailed later in this work) and stored in a database of known-good CandyCodes. When a consumer receives the drug and wishes to verify its authenticity, the consumer uses their cameraphone to take a photo of a CandyCoded pill and uploads this photo to the manufacturer’s server, which performs the same pattern conversion and searches for a CandyCode with a matching or similar pattern in the database. In this example, Consumer 1’s CandyCoded pill has 27 strings (shown in green) that match the strings of a known-good CandyCoded pill in the manufacturer’s database, so the server informs Consumer 1 that their medication is authentic. However, Consumer 2’s CandyCode strings have *no* matches in the database of known-good CandyCodes, so Consumer 2’s drug must not have originated in the manufacturer’s facility and is not authentic, and the server warns Consumer 2 not to consume the medication.

### 2.1 Materials for CandyCodes

Some existing drug products take the form of several small drug-containing spheres enclosed in a transparent capsule. Often these drug-containing spheres are a mixture of two or more colors and are clearly visible from the outside of the capsule. For these particular products, if the drug-containing spheres do not move or shift positions inside the capsule, *it is possible that the existing visible pattern made by the multicolor drug-containing spheres could already function as an intrinsic CandyCode*, and manufacturers could use them as such with *no alterations whatsoever* to their manufacturing process. However, for drug formulations (like the caplets shown in Figure 1) that do *not* contain intrinsic CandyCodes, we can add a CandyCode to the drug using particles.

In principle, any small edible particles with a variety of discernible appearances (different colors, sizes, shapes, etc.) could be used to create a CandyCode on a pill. In this work, I used commercially produced multicolor nonpareils (also known as “sprinkles” and “hundreds and thousands” [11]). Nonpareils come from the manufacturer as a mixture of millimeter-sized candy spheres of different colors. One common variety which was used in this study has eight different sphere colors (each of which is referred to in this work by a single-letter abbreviation as shown): dark blue (D), green (G), light blue (L), orange (O), pink (P), red (R), white (W), and yellow (Y). These nonpareils are available in bulk for around $15 USD per kilogram. At 2.3 milligrams per nonpareil, they cost $0.000 034 5 USD per nonpareil (or 29 000 nonpareils per USD). Additionally, since each nonpareil can have eight different colors, each nonpareil can theoretically encode three binary bits of information (2^3^ = 8), so as a data storage medium, a kilogram of nonpareils could store a theoretical maximum of 1.3 million bits of information.

### 2.2 Adding CandyCodes to drug products

To add CandyCodes to a drug product, a manufacturer needs a method for adhering an adequate number of nonpareils to each pill, tablet, capsule, etc. The method needs to maintain the visibility of nonpareils on the surface of the drug (for photography), be safe for human consumption, and survive mechanical agitation during shipping. As a small-scale pilot test, I used a variety of edible adhesives to affix nonpareils to a typical commercial drug product, 500 mg caplets of acetaminophen/paracetamol (brand name Tylenol). These adhesives included commercial “edible adhesives” (often used to attach edible decorations to cakes), melted edible waxes, and melted sugar. The best results (shown in Figure 1 at right) were obtained using two edible adhesives (details in *Materials and Methods* below). Anecdotally, I found that these “homemade” CandyCoded caplets retained their nonpareil coatings during handling and were actually more pleasant to swallow than plain caplets (confirming Mary Poppins’ classic observation about the relationship between sugar and medicine).

For actual testing, I wanted to use CandyCodes that were more representative of those that would be produced commercially, not my “homemade” versions. I found that commercially produced nonpareil-coated chocolate candies (shown in Figure 1 at left) provide an excellent substitute for actual CandyCoded drug products. These chocolate candies are inexpensive (roughly $0.007 USD per candy when purchased in bulk), already mass-produced by several manufacturers, and roughly the same size as pills and capsules, so they made reasonable and convenient substitutes for actual mass-produced CandyCoded drugs in this study.

### 2.3 Photographing CandyCodes

Once a drug product has received a random coating of nonpareils, the manufacturer must next photograph each CandyCoded pill before packaging. Since many manufacturers already use photographic machine vision systems for quality control purposes, obtaining photographs of CandyCoded drugs should require minimal (if any) alteration to current drug production methods.

To generate a library of CandyCode photos for testing, I randomly selected 120 nonpareil-coated chocolate candies from a one-pound bag and photographed them. I intentionally photographed the candies in groups (12 candies at a time) using a conventional smartphone camera; this resulted in relatively low-resolution images of each CandyCode (shown in Figure 3) as might be expected in a manufacturing facility. These 120 CandyCodes had between 72 and 108 nonpareils per CandyCode, with an average number of 92.6 nonpareils per CandyCode and a median number of 94 nonpareils per CandyCode.

**Figure 3:**
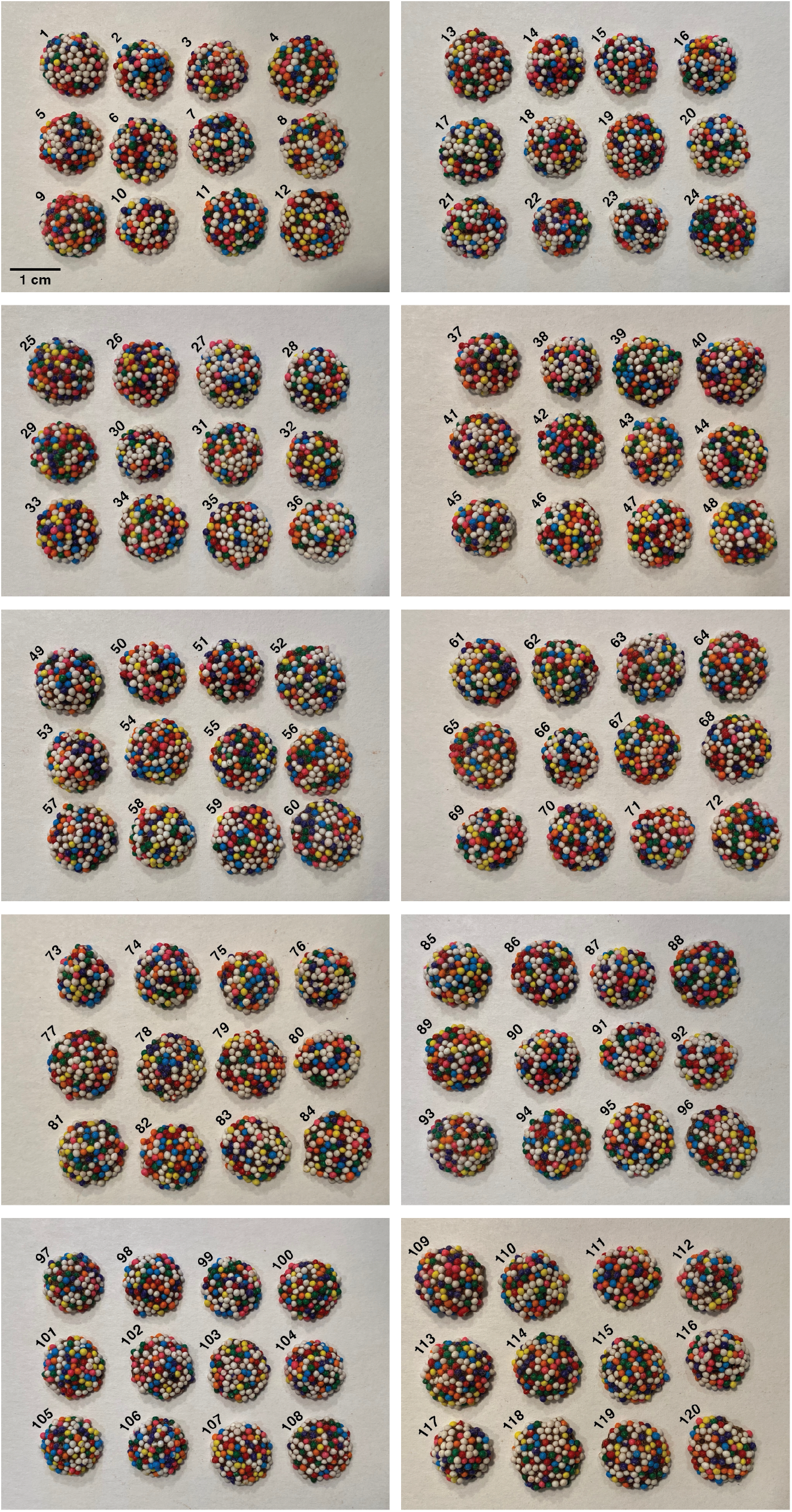
Photographs of 120 candies used to create a test library of CandyCodes. These commercially produced candies were used as proxies for actual CandyCoded medications. Each chocolate candy is coated by the manufacturer with an eight-color mixture of nonpareil spheres; the candies shown here received between 72 and 108 nonpareils each, with an average of 92.6 nonpareils per candy. Each CandyCode photo was converted to a set of strings using the process described in Figure 4 and analyzed to determine its uniqueness. Additionally, three of these CandyCodes (numbers 103, 104, and 109) were selected at random and subjected to one week of rough handling to determine if mechanical wear sustained during shipping could affect the information content of CandyCodes.

### 2.4 Creating a database of known-good CandyCodes

In the next step, the CandyCodes need to be stored in a database for later querying by consumers wishing to verify the authenticity of their CandyCoded medicines. There are many different ways in which this database of known-good CandyCodes could be constructed. Perhaps the most obvious approach would be simply saving the raw CandyCode photos taken by the manufacturer on the production line. When a consumer uploads a photo of a suspect CandyCoded medicine, image similarity algorithms could be used to determine whether there is a match in the database of known-good CandyCode photos. As image similarity algorithms grow more sophisticated, this raw-image-based approach could be a powerful way to build and query a CandyCode database.

Another approach to building a database of known-good CandyCodes would rely upon converting the CandyCode image into standard binary data. This is how one-dimensional barcodes (like EAN/UPC codes [12]) and two-dimensional barcodes (like QR codes [13]) work: a pattern of lines or pixels is decoded to binary data, often interpreted as alphanumeric text. With an average of around 93 nonpareils per CandyCode and 8 different colors per nonpareil, a CandyCode could theoretically encode log_2_ 8^93^ = 279 bits of information. How likely is it that a random 279-bit code could ever be repeated by chance? For comparison, digital “version 4” universally-unique identifiers (UUIDs) commonly used in computing have just 122 random bits [14], and programmers take it for granted that these digital UUIDs will never repeat in normal applications. Thus, a 93-nonpareil CandyCode encoding 279 random bits—*more than double the bits of a digital UUID*—should easily serve as a physical UUID that will *never* be duplicated by random chance.

However, this analysis ignores a crucial difference between barcodes and CandyCodes: barcodes have well-defined structures, like regularly spaced lines or regular grids of pixels, and these structures greatly facilitate their decoding. In other words, each line or pixel in a barcode has a well-defined location, and by reading the lines or pixels at each location, barcodes can easily be decoded and converted into binary information. In contrast, CandyCodes cannot be decoded using a simple barcode-like method because, unlike the lines or pixels in a barcode, the nonpareils in a CandyCode are placed in *random* locations. This random placement is useful—it makes CandyCodes easy to produce and difficult to counterfeit—but it also complicates the process of converting CandyCodes into a binary representation.

That being said, close inspection of CandyCodes like those in Figures 1 and 3 reveals patterns that *can* be used for imparting some local structure to the nonpareil locations, structure that can be useful for decoding CandyCodes. In particular, spherical nonpareils tend to pack onto the flat surface of a pill in a hexagonal pattern, at least in some local areas. This hexagonal pattern lets us identify the nonpareil “neighbors” (usually six of them) that are closest to a given nonpareil. And over larger areas, where the curvature of the pill surface influences the packing of the nonpareils, the patterns start resembling those encountered by mathematicians studying spherical codes and the so-called “Tammes problem” [15]. Local structures like these make it possible to create reasonably robust algorithms for reproducibly identifying “neighborhoods” of nonpareils on the CandyCode. If each neighborhood could be converted to a binary representation, then a CandyCode could be decoded even if it has no overall regular structure.

To test this idea, I wrote a Python program (available in this project’s GitHub repository [16]) that converts a CandyCode photograph into a set of binary text strings. Figure 4 illustrates the individual steps required to convert a specific randomly chosen CandyCode (number 44 from Figure 3) into a set of strings. In Step 1 of Figure 4, the program records the location (in Cartesian coordinates) and color (using the single-letter abbreviations from Section 2.1) of each of the 94 nonpareils visible in the CandyCode photo. These nonpareil coordinates are treated like vertices in a graph. Two vertices in this graph are considered connected by an edge if the corresponding two nonpareils are physically located close enough to be considered “neighbors.” To identify these neighbors, the code calculates the Delaunay triangulation [17] of the nonpareil vertices using an implementation of the Quickhull algorithm [18] in SciPy [19]. In the resulting graph (shown in Step 2 of Figure 4), two vertices connected by an edge are considered neighboring nonpareils.

**Figure 4:**
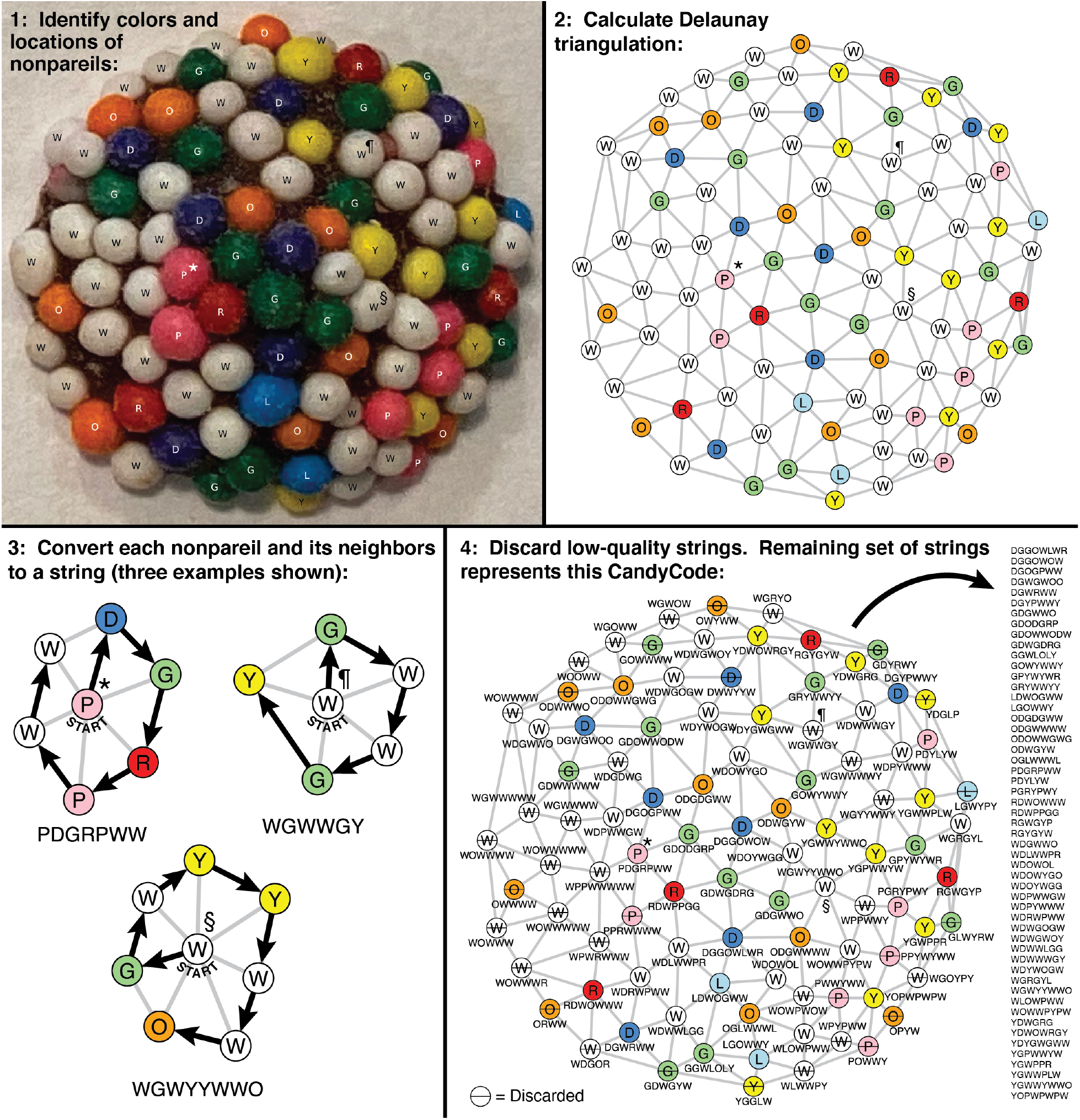
Converting a photograph of a CandyCode (number 44 from Figure 3) into a set of strings suitable for storage and querying in a database. In Step 1, the locations and colors of all visible nonpareils are recorded. In Step 2, the nonpareil locations and colors are interpreted as vertices on a graph, and the Delaunay triangulation [17–19] of these vertices connects neighboring nonpareils via the edges on the graph. Next, for each nonpareil and its immediate neighbors, the colors of each nonpareil and their arrangement around the central nonpareil are used to create a text string. Step 3 shows three example string conversions, for the nonpareil neighborhoods marked with “⋆,” “¶,” and “§” in each step (details in main text). In Step 4, any low-quality strings (*e*.*g*., those containing too-few different colors or located on the edge of the graph) are discarded. The remaining set of 53 strings represents this CandyCode, and this set is saved to the database of known-good CandyCodes. This process is repeated for every CandyCoded pill before leaving the manufacturer. When a consumer photographs a CandyCoded product and submits the photo for analysis, this process is repeated using the consumer’s photo, and the resulting set of strings is compared with the database of known-good string sets. If a known-good CandyCode is found that contains many of the same strings as the user’s CandyCode, then the user likely has an authentic product.

Next, each nonpareil and its immediate neighbors are converted to a text string. For a specific example of this process, find the pink nonpareil marked with “⋆” in the left-of-center region of CandyCode 44 in Figure 4 Step 1. This pink nonpareil has six neighboring nonpareils: two white nonpareils to the left, one dark blue nonpareil above, a green and red nonpareil to the right, and another pink nonpareil below. Now examine the Delaunay triangulation of this CandyCode in Step 2 of Figure 4. The original pink nonpareil’s vertex (again marked “⋆” in Figure 4 Step 2) is connected to six other vertices; these correspond to the pink nonpareil’s six neighboring nonpareils. A closeup showing just the pink nonpareil’s vertex and its six neighboring vertices is shown in Step 3 of Figure 4. To convert this neighborhood of nonpareils to a text string, we start with the single-letter abbreviation of the central nonpareil’s color: P for pink. Next, we sort the single-letter abbreviations of the six connected nonpareils by alphabetical order. The D (dark blue) nonpareil at the top comes first in alphabetical order, so D is added to the string. The remaining neighboring nonpareils are added to the string by moving clockwise around the neighborhood, first to G, then R, then P, then W, and finally the second W. There are no more neighboring nonpareils to add, so the string is complete: PDGRPW W is the string representation of the neighborhood centered on the nonpareil marked “⋆.”

For neighborhoods in which there are two or more surrounding nonpareils of the same color coming first in alphabetical order, the code sorts the two or more *following* vertices by alphabetical order and uses this order to select the next letter for the string. For example, consider the neighborhood centered on the white nonpareil (W) marked with “¶” in Figure 4 Step 3. The string representation of this neighborhood starts with W. Sorting the surrounding nonpareils by alphabetical order reveals that the next letter will be a G, but it is unclear whether this corresponds to the green nonpareil at the top or the second green nonpareil on the bottom. To determine which green nonpareil should come next in the string, the code examines the nonpareils following each green nonpareil moving clockwise. The letter following the top green nonpareil, W, comes earlier in alphabetical order than the letter following the bottom nonpareil, Y. Thus, the top G is chosen as the next letter in the string, the remaining letters are added in the usual clockwise order, and the final string representation for the neighborhood marked “¶” is W GWW GY.

The number of letters in each string is determined by the number of nonpareils in each neighborhood. Many neighborhoods (like the one centered on the nonpareil marked “⋆” in Figure 4) yield seven-letter strings; this is the number predicted by hexagonal packing of spheres as described earlier. However, other string sizes are possible, including smaller strings (like the six-letter string resulting from the neighborhood centered on the nonpareil marked ¶ in Figure 4) and larger strings (like the eight-letter string resulting from the neighborhood centered on the nonpareil marked “§” in Figure 4).

After using this process to convert each nonpareil and its neighbors to a text string as shown in Step 4 of Figure 4, the code discards certain low-quality strings. For example, vertices at the edge of the Delaunay triangulation graph naturally have fewer neighbors, which gives these vertices shorter strings that are more likely to be found repeated in other CandyCodes; discarding these short strings reduces the frequency of “false positives” when searching the database of known-good CandyCodes. Another discard criterion was created in response to an unexpected oddity of the chocolate cand ies used as substitutes for CandyCoded drugs in this study: I found that white nonpareils appear on these candies almost *five times more often* than any other color. In fact, 41.5% of the nonpareils are white, with the remaining seven colors present at only 7.3% to 8.1% each (details in Figure 7). These excess white nonpareils were undoubtedly added intentionally by the manufacturer, but they also have the unfortunate effect of reducing the diversity of letters in the CandyCode strings (nearly half of the letters are W) and again increasing the likelihood of “false positive” strings that appear in multiple CandyCodes. So to eliminate strings with excessive W s, the code discards strings with fewer than four different colors (like the one marked “¶” in Figure 4). In this example, the 94 nonpareils in CandyCode 44 were converted into 94 strings, 41 of which were discarded as low-quality, leaving 53 good strings (shown in the list in Step 4 of Figure 4) that represent this CandyCode in the database of known-good CandyCodes.

This process was repeated for each of the 120 CandyCode images in Figure 3. On average, there were 52.8 good strings per CandyCode. The CandyCode with the smallest number of strings was CandyCode 30, which had just 19 good strings. This is significantly fewer strings than the other CandyCodes, so I examined CandyCode 30 to determine why it stored considerably less information than the other CandyCodes in the test library. I found that CandyCode 30 had an even higher excess of white nonpareils than the other CandyCodes: 55.6% of CandyCode 30’s nonpareils were white, compared to an average of 41.5% white across all 120 CandyCodes. With over half of CandyCode 30’s nonpareils the same color, my algorithm unsurprisingly discarded 44 of the CandyCode’s strings for having fewer than four different colors. Additionally, CandyCode 30 had only 81 total nonpareils, compared to an average of 92.6 nonpareils across all 120 CandyCodes; this gave CandyCode 30 fewer potential strings to begin with. The combination of these two deficiencies—low color diversity and a small nonpareil count—meant that CandyCode 30 had only a third as many strings as the average CandyCode. Both of these deficiencies could easily be eliminated by using equal amounts of each nonpareil color (as discussed later) and ensuring that all CandyCodes receive at least a minimum number of nonpareils.

In contrast, the CandyCode with the largest number of strings was CandyCode 70, which had 73 good strings. This CandyCode had a more diverse selection of colors, with only 36.6% of its nonpareils being white (this is still an excess of white nonpareils, but a smaller excess than the average of 41.5% white). Additionally, CandyCode 70 has a larger-than-average number of nonpareils (101, compared to the average of 92.6). These advantages combine to make CandyCode 70 the most information-rich of all 120 CandyCodes in the test library.

### 2.5 Assessing the uniqueness of CandyCode strings

Normally, once a manufacturer converts a CandyCode photo to a set of strings and adds the set to their database of known-good CandyCodes, a consumer would then take their own photo of the CandyCode and upload it to the manufacturer’s server for analysis and comparison with the database. But before testing a consumer-like interaction with the database, I wanted to determine how how different the database’s CandyCodes were when compared with each other. Specifically, I examined all pairwise combinations of the 120 CandyCodes to determine how many strings the CandyCodes had in common.

Out of the 7140 possible pairwise comparisons among the 120 CandyCodes in the test library:

- 6811 CandyCode pairs (95.4%) had 0 strings in common
- 326 CandyCode pairs (4.56%) had 1 string in common
- 3 CandyCode pairs (0.04%) had 2 strings in common

and no CandyCode pairs had more than 2 strings in common. Stated differently, these results mean that if you choose two CandyCodes at random from the library, over 95% of the time the two codes will have *no* strings in common. Of the small fraction of CandyCode pairs that *do* have strings in common, the vast majority have only a *single string* shared between the CandyCodes. Note that this is a *single shared string* among the *over 100 combined strings* (on average) from both CandyCodes. Finally, an extremely small number of CandyCode pairs (less than one in 2000) have two shared strings. These results show that while repeated strings sometimes appear between two CandyCodes, they are vastly outnumbered by strings that are unique to a single CandyCode, at least for this small-scale test library.

### 2.6 Searching for suspect CandyCodes in the known-good database

Next, to test the experience of consumers using CandyCodes to confirm the authenticity of their medicines, I selected three of the 120 CandyCoded chocolates at random (numbers 103, 104, and 109) and photographed them again (Figure 5A). I then converted these new “suspect” CandyCode photos to string sets as described above, and searched for matching strings in the database of known-good CandyCodes. The results are as follows:

**Figure 5:**
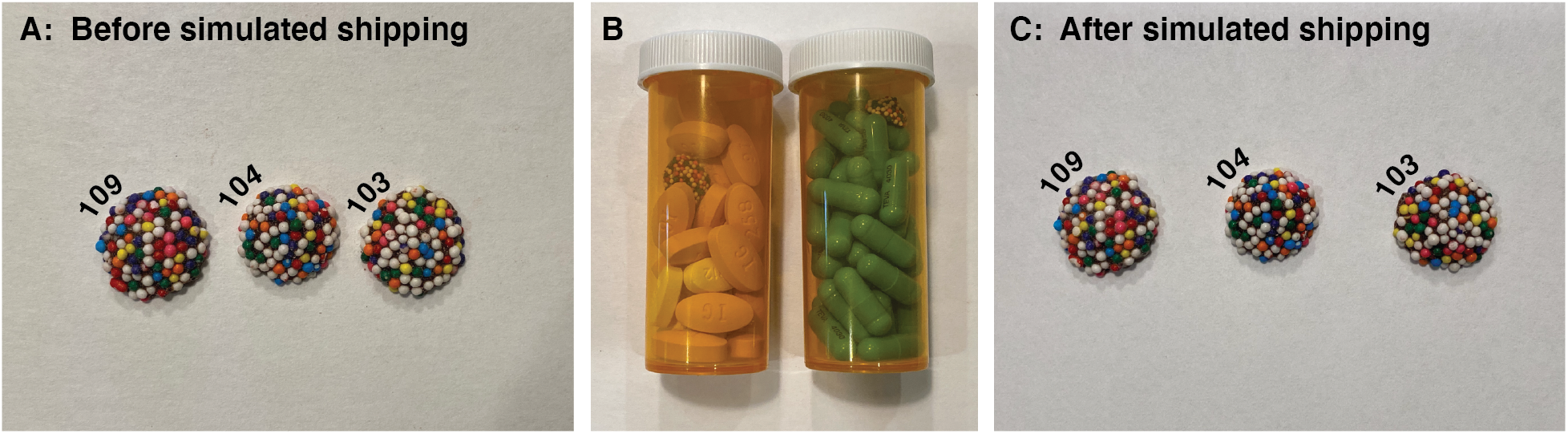
To replicate the experience of a consumer interacting with CandyCoded medications, three CandyCodes from the test library of 120 chocolate-based CandyCodes shown in Figure 3 were selected at random and photographed again **(left)**. These “suspect” CandyCode photos were converted to sets of strings using the process described in Figure 4 and compared with the library of known-good CandyCodes. The matching CandyCodes were found in the library, proving that these “suspect” CandyCodes are indeed authentic (details in the text in Section 2.6). To test whether shipping-induced damage might adversely affect CandyCoded medicines, the same three CandyCodes were added to medicine bottles containing commercial pharmaceutical tablets and capsules **(center)**, and the bottles were then subjected to simulated shipping conditions by tumbling the bottles in a modified rock tumbler nonstop for one week. The CandyCodes were then removed from the bottles and photographed again **(right)**, and the photographs were again converted to sets of strings. Even after undergoing simulated shipping damage, the CandyCodes could still be matched with their entries in the library of known-good CandyCodes to confirm their authenticity (details in the text in Section 2.7).

- **Suspect CandyCode 103** had:
  - 32 strings in common with known-good CandyCode 103
  - 1 string in common with 9 other CandyCodes
  - 0 strings in common with the remaining 110 CandyCodes
- **Suspect CandyCode 104** had:
  - 30 strings in common with known-good CandyCode 104
  - 1 string in common with 7 other CandyCodes
  - 0 strings in common with the remaining 112 CandyCodes
- **Suspect CandyCode 109** had:
  - 23 strings in common with known-good CandyCode 109
  - 2 strings in common with 1 other CandyCode
  - 1 string in common with 3 other CandyCodes
  - 0 strings in common with the remaining 115 CandyCodes.

In other words, the suspect CandyCode photos had an average of *28 strings* in common with their corresponding photos in the known-good database; this proves that the suspect CandyCodes have matches in the known-good database and are therefore authentic. In addition, the suspect CandyCodes had zero strings in common with most of the non-matching CandyCodes in the database, and never had more than 2 strings in common with non-matching CandyCodes.

While there are many strings in common between the “suspect” photo and the “known-good” photo of the same CandyCode, the string sets are not identical between the pairs of photos. Specifically, CandyCode 103’s photos had 32 matching strings out of 53 (or 60% of the strings matching), CandyCode 104 had 30 matching strings out of 49 (61% matching), and CandyCode 109 had 23 matching strings out of 57 (40% matching). Stated differently, *two photos of the same CandyCode do not always result in the exact same set of strings*. This can be attributed to the process used to convert CandyCode photos to string sets. In particular, the Delaunay triangulation (which is used to identify neighboring nonpareils) is very sensitive to the angles formed between nearby nonpareils, angles that could vary in different photographs taken from slightly different vantage points. But even though different photos of the same CandyCode have only about half of their strings in common, this is still more than enough shared strings to determine that the CandyCodes are the same.

### 2.7 Verifying CandyCodes after simulated shipping conditions

CandyCodes use nonpareils attached to a pill to impart a unique identifier to the pill. These attached nonpareils make CandyCoded medicines easier to produce compared to anti-counterfeiting measures that require reformulation of a drug product. But if the nonpareils are not firmly affixed to the medication, they could become detached during the mechanical agitation that might accompany shipping and distributing the product. If enough nonpareils are lost, a consumer’s CandyCoded medication might no longer match the manufacturer’s record of that CandyCode, and the consumer would be told that the product is fraudulent when in fact it is authentic.

To examine the likelihood of a damaged CandyCode providing a consumer with incorrect information, I took the same three chocolate-based CandyCodes examined in the previous section (numbers 103, 104, and 109) and subjected them to simulated shipping conditions. Each CandyCode was placed in its own standard pill bottle accompanied by assorted commercial medications (tablets and capsules) as shown in Figure 5B. To intentionally maximize the mechanical stresses on the CandyCodes, the bottles were not fully filled and the empty space was not filled with any cotton batting, leaving the CandyCodes free to rattle and collide with the pills. The pill bottles were then placed into a modified rock tumbler and subjected to constant tumbling for one full week. The tumbler inverted the pill bottles once every two seconds, or over 300 000 inversions over the course of the experiment. After a week of tumbling, the CandyCodes were removed from the bottles and photographed again (Figure 5C), and each CandyCode photo was converted to a set of strings as described above. Finally, I compared these post-tumbling CandyCode strings to the original library of 120 CandyCodes to determine if the CandyCodes that underwent simulated shipping could still be identified in the library.

The results show that simulated shipping conditions had little effect on the appearance or function of the CandyCodes. Close comparison of the before and after photos in Figure 5 show that a couple of CandyCode 109’s nonpareils were chipped during a week of simulated shipping, but the colors of these nonpareils were still recognizable. Converting the photos of the worn CandyCodes to string sets and comparing them with the original 120-CandyCode library had these results:

- **Post-shipping CandyCode 103** had:
  - 27 strings in common with known-good CandyCode 103
  - 1 string in common with 9 other CandyCodes
  - 0 strings in common with the remaining 110 CandyCodes
- **Post-shipping CandyCode 104** had:
  - 21 strings in common with known-good CandyCode 104
  - 1 string in common with 8 other CandyCodes
  - 0 strings in common with the remaining 111 CandyCodes
- **Post-shipping CandyCode 109** had:
  - 27 strings in common with known-good CandyCode 109
  - 1 string in common with 2 other CandyCodes
  - 0 strings in common with the remaining 117 CandyCodes.

Comparing these results to the pre-shipping results for the same CandyCodes in the previous section, we see that after shipping, two CandyCodes had small decreases in the number of strings shared with the same CandyCode in the known-good database (CandyCode 103’s string matches dropped from 32 to 27, and CandyCode 104’s string matches dropped from 30 to 21), and one CandyCode actually had a small *increase* in the number of matching strings (CandyCode 109’s matches rose from 23 to 27). In light of these seemingly random changes, I attributed the changes to variations in the Delaunay triangulation from different photos of the same CandyCode as described earlier, not effects of the simulated shipping.

### 2.8 Simulating CandyCode libraries

The experiments described above used a test library of just 120 CandyCodes. In real-world applications involving a commercial pharmaceutical, the library of CandyCodes would be far larger. To explore larger CandyCode libraries without using enormous numbers of chocolate candies, I created a Python-based simulator (available on this project’s GitHub repository [16]) that generates CandyCodes on a computer. The simulator replicates real CandyCode production by randomly placing small circles of different colors (the nonpareils) onto a larger circular area (the pill). The user can also specify the number of nonpareils per CandyCode, the number and probability of nonpareil colors, and the total number of CandyCodes to generate. The simulated CandyCodes are then subjected to the same analysis as the test library above, including conversion to string sets and analysis for matching strings between pairs of CandyCodes. Finally, this entire process was repeated 100 times for each set of simulation parameters, and the results were collected for statistical analysis.

To test the CandyCode simulator, I first used it to replicate the results of the experimental 120-CandyCode test library described above. Each of the 120 simulated CandyCodes had 94 nonpareils (the median number of nonpareils per CandyCode in the test library). The simulated nonpareil color frequency also matched the test library, with white nonpareils appearing five times more frequently than the other individual colors. After simulating and analyzing the 120-CandyCode library 100 separate times, I found that the largest number of strings shared between any two different CandyCodes in a library was 2.26 on average. This result is consistent with the experiments in Section 2.5 above, which found no more than 2 shared strings among the 120 real (chocolate-based) CandyCodes.

Having replicated the experimental results using the simulator, I then simulated CandyCode libraries with sizes ranging from 1 to 100 000 CandyCodes per library (and still simulating each library size 100 times for statistical analysis). The results are shown in red in Figure 6A, which plots the largest number of strings shared between any two different CandyCodes (on a linear scale) versus the total number of CandyCodes in the library (on a logarithmic scale). Not surprisingly, as the size of the CandyCode library increases, shared strings between the CandyCodes become more common. But the increase in shared strings follows a logarithmic function of library size, and least-squares fitting shows that increasing the library size by a factor of 10 only increases the largest number of shared strings by a factor of 1.2. So the largest library I simulated, 100 000 CandyCodes, would only be expected to have about 5.5 strings shared between any two different CandyCodes in the library. Since the smallest number of strings shared between two photos of the *same* CandyCode was 21 (determined in Section 2.7 above), it would be trivial to distinguish a correct CandyCode with 21 or more matching strings from incorrect CandyCodes with only 5 or 6 matching strings. So even with an excess of white nonpareils, these simulations suggest that the eight-color CandyCodes could support at least 100 000 different and distinguishable CandyCodes.

**Figure 6:**
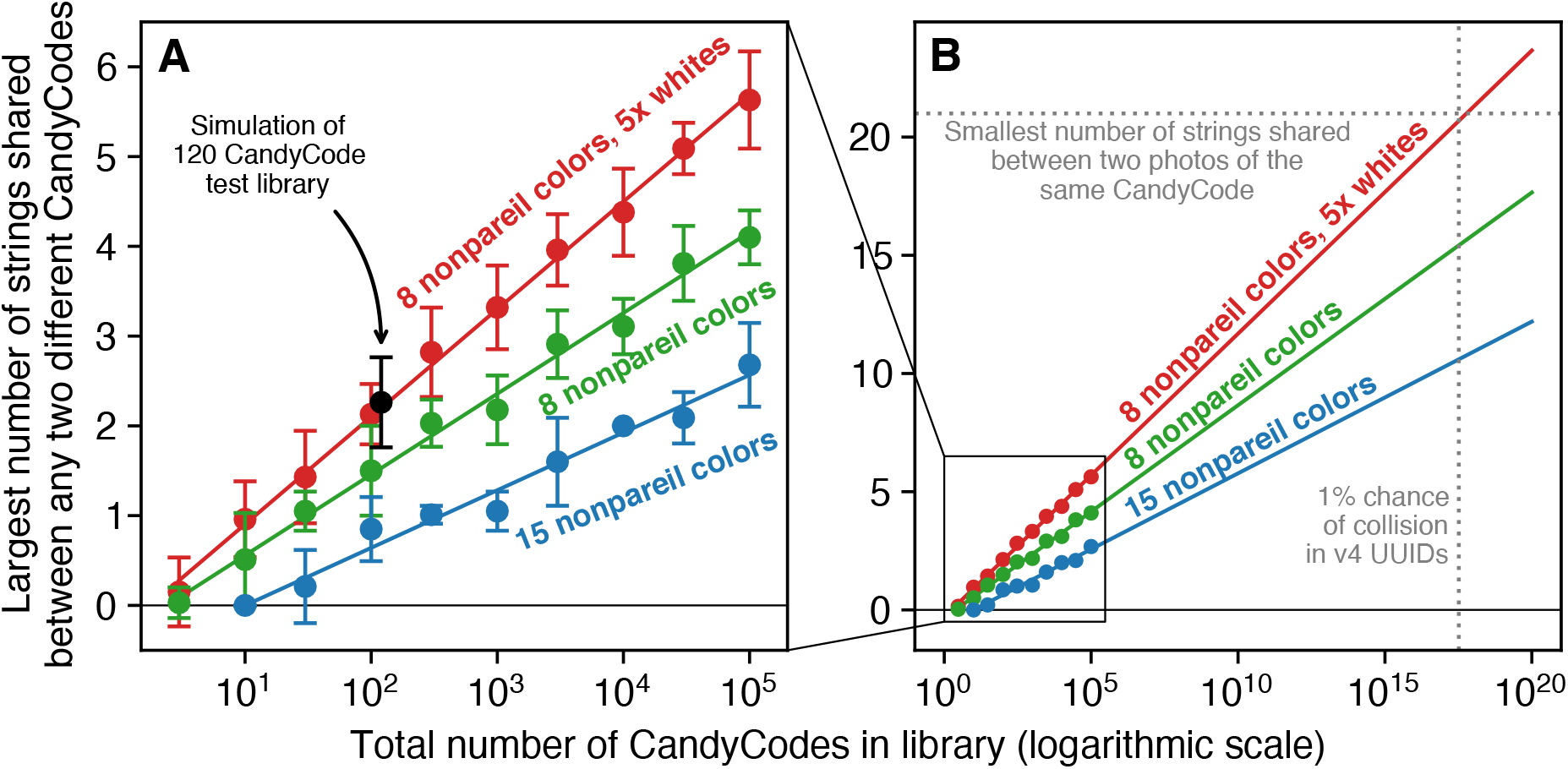
Results from analyzing simulated CandyCode libraries of different designs and sizes. Each point represents the average of 100 separate simulations of the CandyCode library, with error bars ±1 standard deviation. **(A)** A simulation of Figure 3’s 120-CandyCode test library with eight colors and a 5× excess of white nonpareils **(black point)** predicts an maximum of 2.26 strings shared between any two different CandyCodes; this is in good agreement with the experimentally observed value of 2 maximum shared strings for this library. Simulating larger libraries of eight-color CandyCodes with excess white nonpareils **(red points)** reveals a logarithmic relationship with library size **(red line)**; increasing the library size by a factor of 10 increases the largest number of shared strings by a factor of 1.2. Using CandyCodes with eight equal-probability colors **(green points and line)** reduces the number of strings shared between any two different Codes, so that the same 10× increase in library size only increases the largest number of shared strings by a factor of 0.9. Finally, using CandyCodes with 15 equal-probability colors **(blue points and line)** further reduces the number of shared strings, so that the same 10 ×increase in library size only increases the largest number of shared strings by a factor of 0.6. **(B)** By extrapolating these results to a CandyCode library size of 10^17^ (the number of “version 4” digital UUIDs [14] that must be generated before the odds of having a repeated UUID reach 1% [20]), we can predict that a CandyCode library of that size using eight colors and a 5 × excess of white nonpareils would have up to 21 strings shared between any two CandyCodes; this equals the smallest number of strings shared between two photos of the *same* CandyCode in my experiments, so CandyCodes from this library would probably *not* be suitable for use as universally unique indentifiers. However, a similarly-sized library using eight equal-probability colors is predicted to have only up to 15 strings shared, and a library using 15 equal-probability colors should have only up to 10 strings shared. These projections suggest that CandyCodes from these libraries *could* be used as universally-unique identifiers, and each CandyCode in the library could be distinguished from the 10^17^ other CandyCodes in the library.

Libraries with more than 100 000 CandyCodes were too large to generate and analyze in a reasonable amount of computing time, but by extrapolating the simulation results, we can try to predict the feasibility of much larger CandyCode libraries. The red line in the zoomed-out plot in Figure 6B shows that for eight-color nonpareils with a 5× excess of whites, the predicted largest number of strings shared between any two different CandyCodes in the library will exceed 21 after about 10^17^ CandyCodes are generated. I chose 10^17^ as a target number of CandyCodes because it is roughly equal to the number of “version 4” digital UUIDs [14] that can be generated on a computer before the probability of a duplicated UUID reaches 1% [20]. These digital UUIDs are routinely treated as universally unique in computing, so if 10^17^ CandyCodes can be distinguished from each other, we can assume that the CandyCodes are also universally unique and could be used to uniquely identify one pill out of 10^17^ pills (this enormous number equals *41 million pills for every person on Earth*).

In the case of CandyCodes with eight-color nonpareils and a 5× excess of white nonpareils, the prediction in red in Figure 6B suggests that a library of 10^17^ CandyCodes is likely *not* suitable for use as a unique identifiers. Some mismatched CandyCodes in this library will still have up to 21 strings in common, and it would be impossible to distinguish these mismatched codes from correctly-matched CandyCodes that may have as few as 21 strings in common. So while the commercially produced candies used in this study could probably be uniquely identified in libraries of ∼10^10^ CandyCodes (which may be more than large enough for many pharmaceutical applications), they probably cannot be considered universally unique at the same level as digital UUIDs.

### 2.9 Simulating CandyCode libraries with unbiased nonpareil colors

While the commercially-produced candies we used as test CandyCodes had a 5× excess of white nonpareils, it would be trivial for a manufacturer to use equal amounts of each color of nonpareil. I hypothesized that by using equal amounts of each nonpareil color, fewer strings would be discarded for having too-few unique colors, and the resulting CandyCodes could still be distinguishable even in larger libraries.

To test this hypothesis, I repeated the eight-color CandyCode simulations described in the previous section, but with each color having the same (12.5%) probability as shown in Figure 7B. The simulation results shown in green in Figure 6 confirm that using equal amounts of each nonpareil color does reduce the number of strings shared between different CandyCodes. In this model, increasing the size of an equal-colored CandyCode library by a factor of 10 only increases the largest number of strings shared between any two different CandyCodes by a factor of 0.9 (compared to a factor of 1.2 for CandyCodes with 5× excess whites). This means that larger CandyCode libraries are feasible using equal amounts of nonpareil colors: the extrapolation in Figure 6B predicts that up to 15 strings would be shared between different CandyCodes in a universally-unique library of 10^17^ CandyCodes. This number is lower than 21, the smallest number of strings shared between photos of the *same* CandyCode observed in my experiments, so this projection suggests that an eight-color CandyCode with equal color probabilities *could* serve as a universally unique identifier, with CandyCodes still distinguishable from each other even after 10^17^ of them have been made.

**Figure 7:**
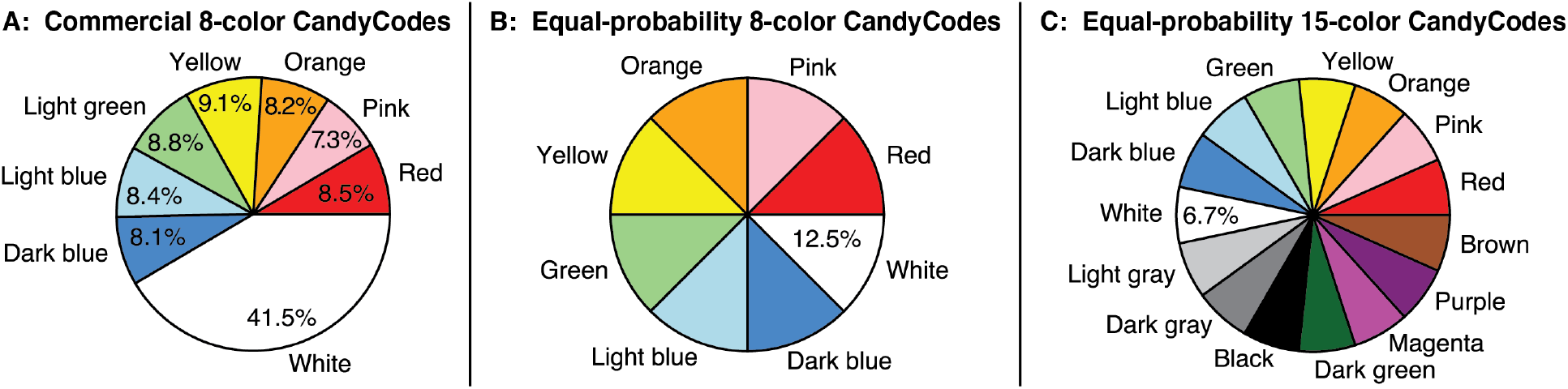
Probabilities of nonpareil colors in the three CandyCode systems analyzed in this work: **(A)** the commercial eight-color chocolate candies with excess white nonpareils, used in the 120-CandyCode test library in Sections 2.1–2.7 and simulated in Section 2.8; **(B)** the proposed eight-color CandyCode with equal probabilities for each color, simulated in Section 2.9; and **(C)** the proposed fifteen-color CandyCode system with equal probabilities for each color, simulated in Section 2.10.

### 2.10 Simulating CandyCode libraries with additional colors

The experiments and simulations described above all use eight colors of nonpareils because many commercial products with nonpareils have eight colors. However, nothing fundamentally limits CandyCodes to using just eight colors. In principle, one could use as many different nonpareil colors as can be reliably discriminated using a camera. While modern smartphone cameras can distinguish a massive number of different colors, CandyCodes should probably limited to a subset of colors that can be reliably distinguished even under sub-optimal lighting and other adverse conditions.

I theorized that I could add seven additional colors to the original eight colors to create a set of 15 nonpareil colors that could still be reliably distinguished in a photograph. The seven new colors—black, dark gray, light gray, dark green, magenta, purple, and brown—are shown along with the original eight colors in Figure 7C. With so many colors of nonpareils, the odds of discarding a text string due to too-few colors become very small, and even-larger libraries of CandyCodes should be feasible.

The 15-color CandyCode model was then subjected to the same simulation and analysis as the previous models. The results, shown in blue in Figure 6, confirm that adding additional colors of nonpareils further reduces the number of strings shared between different CandyCodes. In a 15-color CandyCode library, increasing the number of CandyCodes by a factor of 10 only increases the largest number of strings shared between any two different CandyCodes by a factor of 0.6; this means that a 15-color CandyCode library should have roughly *half* as many strings shared between different CandyCodes as an equivalently sized library containing 8-color CandyCodes with 5× excess whites. Consequently, even larger libraries are feasible using 15 nonpareil colors, and Figure 6B predicts that a library containing 10^17^ 15-color CandyCodes should have a maximum of only about 10 strings shared between different CandyCodes. This number is well below the minimum observed number of strings shared between photos of the *same* CandyCode in my experiments, so determining whether a given 15-color CandyCode exists in a library of 10^17^ CandyCodes should be quite feasible (though further experimental testing would be necessary to confirm that common cellphone cameras can indeed reliably distinguish 15 different colors of nonpareils).

## 3 Discussion

This work demonstrates the feasibility of using random patterns of multicolored particles as universal identifiers. The specific implementation of CandyCodes described here is functional, but there is definitely room for improvement. In this section, I conclude by commenting on possible enhancements and applications for this technique in the future.

Perhaps most significantly, the process I developed for storing known-good CandyCodes in a searchable database could be dramatically improved. Converting each CandyCode photograph into a set of strings, as was done here, makes for a compact and easily searchable database, and my projections suggest that this string-based approach can scale to UUID-sized libraries (∼10^17^ CandyCodes), but text conversion also discards a great deal of useful information contained in the CandyCode photographs. Storing known-good CandyCodes as raw photos instead of strings, and then using existing image search/comparison algorithms to quantify CandyCode similarity and find matches, could unlock much more of the information content of CandyCodes and further reduce the risk of erroneously classifying a fraudulent product as authentic (or vice versa). Recent breakthroughs in the fields of machine learning and computer vision could prove especially useful in CandyCode analysis.

Additionally, modifications to the CandyCode design shown here could impart additional robustness or error-resistance to CandyCodes. For example, I used identically-sized and -shaped spherical nonpareils in this work because they are readily available, but CandyCodes are not limited to using these exact nonpareils. Using two or three different *sizes* of nonpareils would double or triple the number of distinguishable particles used to create the CandyCode. Similarly, using different *shapes* of nonpareils (like triangles, rods, squares, and so on) would add additional uniqueness to the CandyCodes. For CandyCodes applied to noncircular forms of medications (like the oblong caplets shown in Figure 1), the shape of the medication could be used to impose an overall orientation or alignment on the CandyCode, simplifying its analysis and comparison with other CandyCodes.

Finally, while this work focused on using CandyCodes to combat fraudulent pharmaceuticals, many other products are routinely counterfeited and passed as authentic. In 2019, the US Customs and Border Protection agency seized over $1.5 billion USD worth of counterfeit goods [21]. Cosmetics, perfumes, wine, spirits, apparel, and accessories are some of the most-frequently counterfeited types of goods. To defend their genuine products, a manufacturer could simply apply a drop of glue to the cap of a perfume or beverage bottle, then randomly apply colored particles on the glue during the manufacturing process, forming a universally-unique CandyCode on the product (as shown in Figure 8). Similarly, the tag on a dress, handbag, or wallet could receive a sticker that has been dusted with multicolored glitter. In each of these cases, a consumer would be encouraged to use their smartphone to snap a photo of the CandyCode and submit it to the manufacturer for confirmation before purchasing the product. Since CandyCodes are simple to make but difficult to duplicate, they can add value and defend against fraud in a wide variety of different products.

**Figure 8:**
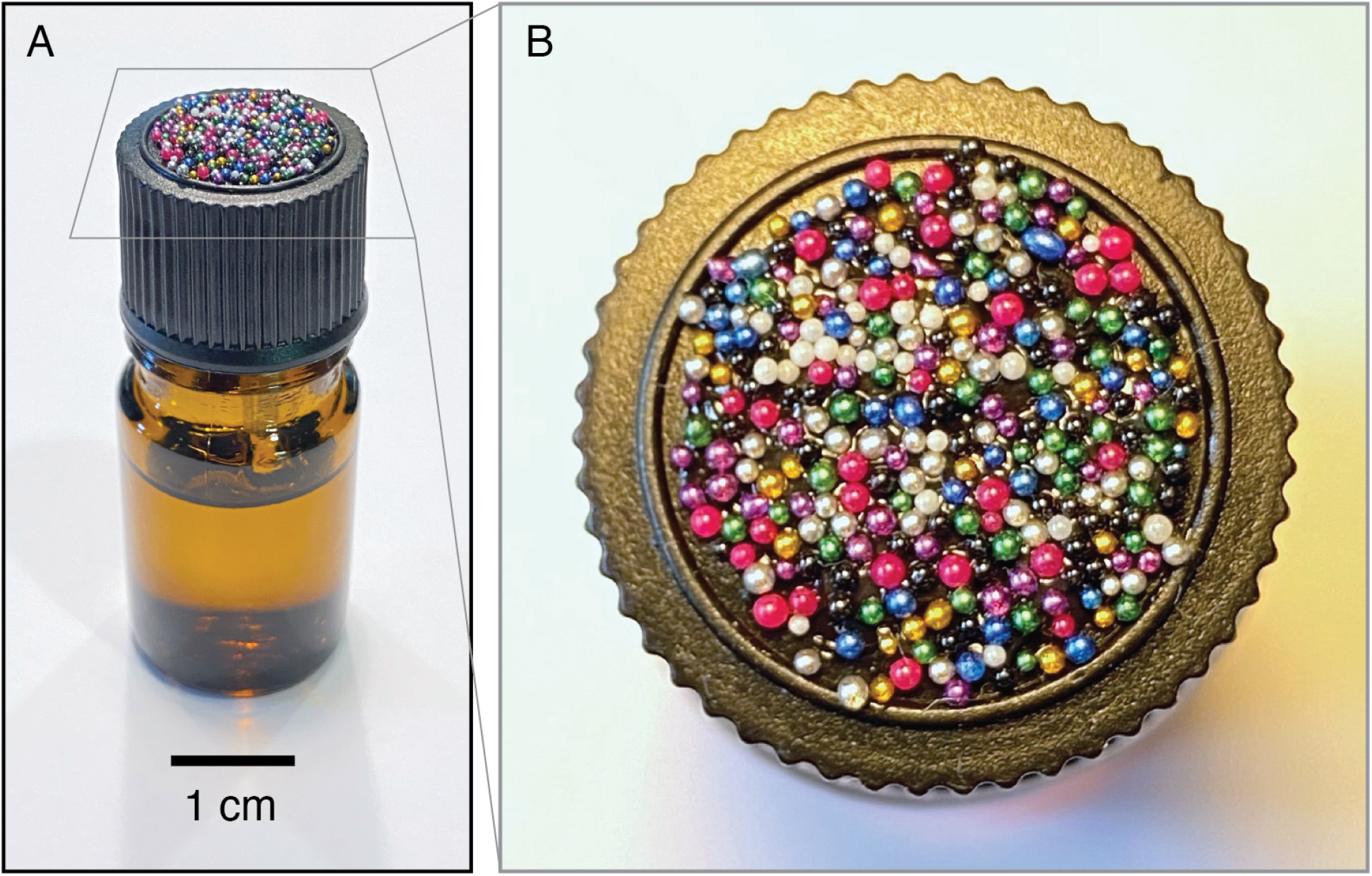
By applying adhesive to the cap of a small bottle and then pressing the cap into a bed of multicolored plastic spheres, the bottle receives a universally unique CandyCode that can subsequently be photographed and used to verify the authenticity of the bottle’s contents (assuming that the seal is intact). This CandyCode is only about a centimeter in size but contains nearly 300 easily-distinguishable microbeads; this is almost three times the number of nonpareils per CandyCode used in this work, meaning that this bottle’s CandyCode is universally unique and will never be duplicated by random chance.

## 4 Materials and methods

In principle, any suitably sized and distinguishable particles could be used to make CandyCodes. The materials used in this demonstration were chosen for their commercial availability and low cost. The bulk nonpareils shown in Figure 1 were “Over the Top” brand (manufactured by Topco Associates, Elk Grove Village, IL, USA). These nonpareils were affixed to caplets of a commercial drug (500 mg caplets of acetaminophen/paracetamol, brand name Tylenol; manufactured by McNeil Consumer Healthcare, Fort Washington, PA, USA) using edible adhesive. After testing several different types of edible adhesive, I found two brands that gave the best results: “Sugarcraft Essentials” edible glue (containing water, preservative, acetic acid, and carboxymethyl cellulose; manufactured by Rainbow Dust Colors, Cuerden Green Mill, UK), and “Dab-N-Hold” edible adhesive (containing water, tapioca dextrin, dextrose, citric acid, xanthan gum, propylene glycol, sodium benzoate, and artificial flavors; manufactured by Wilton Industries, Naperville, IL, USA). The bulk nonpareil-coated chocolate candies shown in Figure 1 and used in Figures 3–5 were purchased from “Nuts.com” (Cranford, NJ, USA). The ∼850 µm diameter multicolored plastic spheres shown in Figure 8 were “Slime Metallic Microbeads,” produced for arts and crafts applications by Maddie Rae’s (Trumbull, CT, USA); they were adhered to the cap of a small perfume bottle using silicone adhesive (GE brand; Momentive Performance Materials, Huntersville, NC, USA). All CandyCodes were photographed using a conventional smartphone camera (iPhone 11 Pro; Apple, Cupertino, CA, USA) under ordinary room lighting conditions. A set of custom Python programs (available at this project’s GitHub repository [16]) was used to analyze the library of 120 chocolate-candy-based CandyCodes shown in Figure 3 and simulate the CandyCode libraries in Figure 6.

## Data Availability

All data referred to in the manuscript is available at the associated GitHub repository.

https://github.com/groverlab/candycodes

## Notes

Supported by a grant from the Bill and Melinda Gates Foundation (OPP1191214).

### Competing Interest Statement

The authors have declared no competing interest.

### Funding Statement

This work was supported by a grant from the Bill and Melinda Gates Foundation (OPP1191214).

### Author Declarations

This research did not involve human subjects or any other aspects requiring institutional approval.

### Summary of Updates

One additional paragraph has been added to the Introduction that describes relevant prior research in the field of “Physical Unclonable Functions” (PUFs). The rest of the manuscript is unchanged.

## References

[1] World Health Organization. WHO Global Surveillance and Monitoring System for substandard and falsified medical products. Geneva, 2017.

[2] SofMat Anti-Counterfeiting Solutions. https://sofmat.com/sequential-marking/, 2021.

[3] Minli You, Min Lin, Shurui Wang, Xuemin Wang, Ge Zhang, Yuan Hong, Yuqing Dong, Guorui Jin, and Feng Xu. Three-dimensional quick response code based on inkjet printing of upconversion fluorescent nanoparticles for drug anti-counterfeiting. Nanoscale, 8(19):10096–104, May 2016.

[4] Sangkwon Han, Hyung Jong Bae, Junhoi Kim, Sunghwan Shin, Sung-Eun Choi, Sung Hoon Lee, Sunghoon Kwon, and Wook Park. Lithographically encoded polymer microtaggant using high-capacity and error-correctable QR code for anti-counterfeiting of drugs. Adv Mater, 24(44):5924–9, Nov 2012.

[5] Jie Fei and Ran Liu. Drug-laden 3d biodegradable label using qr code for anti-counterfeiting of drugs. Mater Sci Eng C Mater Biol Appl, 63:657–62, Jun 2016.

[6] Maren Preis, Joerg Breitkreutz, and Niklas Sandler. Perspective: Concepts of printing technologies for oral film formulations. Int J Pharm, 494(2):578–584, Oct 2015.

[7] Magnus Edinger, Daniel Bar-Shalom, Niklas Sandler, Jukka Rantanen, and Natalja Genina. QR encoded smart oral dosage forms by inkjet printing. Int J Pharm, 536(1):138–145, Jan 2018.

[8] Ravikanth Pappu, Ben Recht, Jason Taylor, and Neil Gershenfeld. Physical one-way functions. Science, 297(5589):2026–2030, 2002.

[9] Riikka Arppe and Thomas Just Sørensen. Physical unclonable functions generated through chemical methods for anti-counterfeiting. Nature Reviews Chemistry, 1(4):1–13, 2017.

[10] Jung Woo Leem, Min Seok Kim, Seung Ho Choi, Seong-Ryul Kim, Seong-Wan Kim, Young Min Song, Robert J Young, and Young L Kim. Edible unclonable functions. Nat Commun, 11(1):328, 01 2020.

[11] Tim Richardson. Sweets: A History of Candy. Bloomsbury, New York, 2002.

[12] ISO. 15420:2009 Information technology: Automatic identification and data capture techniques: EAN/UPC bar code symbology specification. Technical report, 2009.

[13] ISO. 18004:2015 Information technology: Automatic identification and data capture techniques: QR Code bar code symbology specification. Technical report, 2015.

[14] Paul J. Leach, Rich Salz, and Michael H. Mealling. A Universally Unique IDentifier (UUID) URN Namespace. RFC 4122, July 2005.

[15] P. Delsarte, J.M. Goethals, and J.J. Seidel. Spherical codes and designs. In D.G. Corneil and R. Mathon, editors, Geometry and Combinatorics, pages 68–93. Academic Press, 1991.

[16] William H. Grover. Candycodes: Simple universally unique edible identifiers for confirming the authenticity of pharmaceuticals. https://github.com/groverlab/candycodes, 2021.

[17] B. N. Delaunay. Sur la sphère vide. Bull. Acad. Sci. URSS, 1934(6):793–800, 1934.

[18] C Bradford Barber, David P Dobkin, and Hannu Huhdanpaa. The Quickhull algorithm for convex hulls. ACM Transactions on Mathematical Software (TOMS), 22(4):469–483, 1996.

[19] Pauli Virtanen, Ralf Gommers, Travis E. Oliphant, Matt Haberland, Tyler Reddy, David Cournapeau, Evgeni Burovski, Pearu Peterson, Warren Weckesser, Jonathan Bright, Stéfan J. van der Walt, Matthew Brett, Joshua Wilson, K. Jarrod Millman, Nikolay Mayorov, Andrew R. J. Nelson, Eric Jones, Robert Kern, Eric Larson, C J Carey, İlhan Polat, Yu Feng, Eric W. Moore, Jake VanderPlas, Denis Laxalde, Josef Perktold, Robert Cimrman, Ian Henriksen, E. A. Quintero, Charles R. Harris, Anne M. Archibald, Antônio H. Ribeiro, Fabian Pedregosa, Paul van Mulbregt, and SciPy 1.0 Contributors. SciPy 1.0: Fundamental Algorithms for Scientific Computing in Python. Nature Methods, 17:261–272, 2020.

[20] Frank H. Mathis. A generalized birthday problem. SIAM Review, 33(2):265–270, 1991.

[21] US Customs and Border Protection. Intellectual property rights: Fiscal year 2019 seizure statistics. https://www.cbp.gov/document/report/fy-2019-ipr-seizure-statistics, 2019.

